# ALeRT-COVID: Attentive Lockdown-awaRe Transfer Learning for Predicting COVID-19 Pandemics in Different Countries

**DOI:** 10.1101/2020.07.09.20149831

**Authors:** Yingxue Li, Wenxiao Jia, Junmei Wang, Jianying Guo, Qin Liu, Xiang Li, Guotong Xie, Fei Wang

## Abstract

Countries across the world are in different stages of COVID-19 trajectory, among which many have implemented the lockdown measures to prevent its spread. Although the lockdown is effective in such prevention, it may put the economy into a depression. Predicting the epidemic progression with government switching the lockdown on or off is critical. We propose a transfer learning approach called ALeRT-COVID using attention-based recurrent neural network (RNN) architecture to predict the epidemic trends for different countries. A source model was trained on the pre-defined source countries and then transferred to each target country. The lockdown measure was introduced to our model as a predictor and the attention mechanism was utilized to learn the different contributions of the confirmed cases in the past days to the future trend. Results demonstrated that the transfer learning strategy is helpful especially for early-stage countries. By introducing the lockdown predictor and the attention mechanism, ALeRT-COVID showed a significant improvement on the prediction performance. We predicted the confirmed cases in one week when extending and easing lockdown separately. Results showed the lockdown measures is still necessary for a number of countries. We expect our research can help different countries to make better decisions on the lockdown measures.

## Introduction

The world is going through the COVID-19 pandemic. As of June 21, the cumulative case number has reached 8,952,419 and the total death number is up to 468, 392 around the globe^1^. The COVID-19 pandemic hit China in the late December of 2019 and then spread to countries in Europe and America^2,3^. Now it is accelerating in densely populated developing countries in Asia, Africa and South America. The current COVID-19 epidemiological situation alterts the world to be prepared for the global health crisis. Many countries have implemented lockdown measures to control the epidemic progression^4–6^ at different time (Fig. 1). Specific lockdown^7–10^ measures adopted included closing schools, churches, bars and other social venues, limiting travel and public gatherings and even shutting down factories and business and staying at home. These interventions helped these countries reduce case numbers and mortality rate at different levels. Under the economic pressure^11,12^, some countries have lifted the lockdown measures and some are planing to. However, the epidemic progression may rebound once these controls are lifted. Therefore, forecasting the epidemic progression when the interventions start to be implemented or lifted is of crutial importance. Such prediction can provide valuable information for better understanding the current situation and help policy makers and health authorities make appropriate plans to manage the country. However, the challenge lies in making predictions for the countries in the early-stage of the COVID-19 pandamic, which are with too scarce data to train an accurate prediction model.

**Figure 1.**
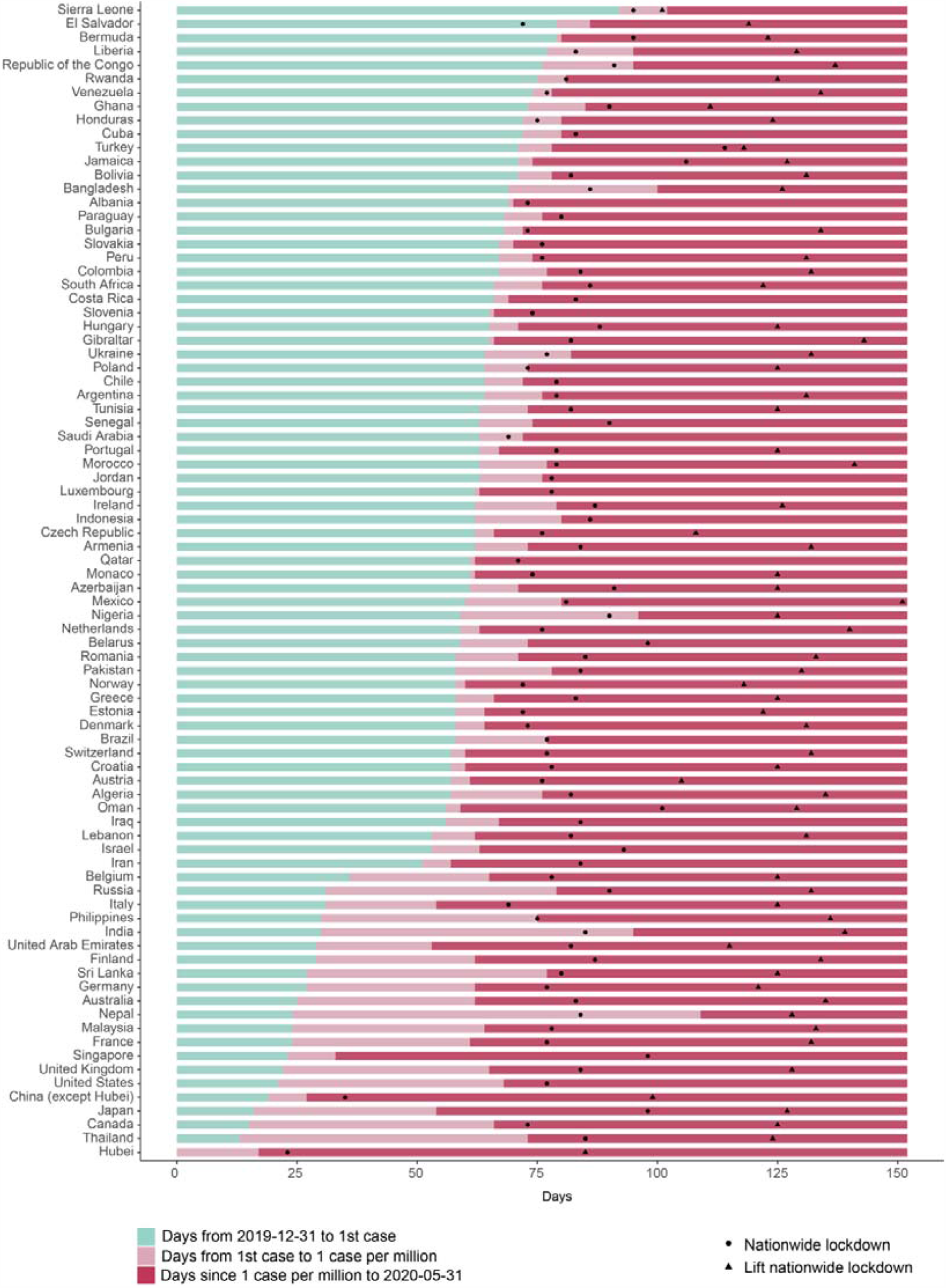
Timeline of COVID-19 dispersion of different countries and government responses. The start point of the X-axis is the date when China reported a cluster of COVID-19 cases in Wuhan, Hubei Province (2019-12-31). The length of the green bar denotes the days from 2019/12/31 to the date of the first reported cases in each specific country. The length of the light pink bar denotes the days from the 1^st^ case to reaching 1 case per million in each country. The dark pink bar denotes the days from the date of reaching 1 case per million to May 31. The x-coordinate of the black dot means the days from the start point to the start date of the lockdown measure. The x-coordinate of the black triangle means the days from the start point to the end date of the lockdown measure. Among the countries which were still under lockdown before May 31 (no triangle presents in the corresponding panel), Indonesia, Singapore and United States had announced their lockdown end date as July 31, June 1 and June 1 respectively.

There have been existing studies on predicting the spread of COVID-19^13–17^. Traditional epidemiology models, such as susceptible–infected (SI)^17,18^, SI recovered (SIR)^19^, and susceptible–exposed–infected–recovered (SEIR)^20–23^, analyze the infection rate based on the dynamic change in the number of infections and subsequently predict the spread and development trend of the epidemic. However, these models made strong assumptions on the infection dynamics and are appropriate to predict the long-term trend of the epidemic. Time series analysis models, such as autoregressive integrated moving average (ARIMA), have also been applied to predict the epidemic trends. ARIMA model can cover a wide range of patterns stationary to non-stationary and seasonal (periodic) time series, but their prediction performance is limited by their reliance on the prior knowledge of model parameters or inherent time-lags. Furthermore, these models do not account for additional factors that can impact the development of infectious diseases^24^. Recently, deep learning algorithms, such as recurrent neural network (RNN), have also been aplied to predict the epidemic progression^15,21^. These models are completely data-driven and do not reply on any prior assumptions. Although they have been demonstrated to be able to achieve prediction performance, they so require a large number of training samples due to their complex architectures^25,26^. The countries at early stage of COVID-19 only have limited data points, which makes it difficult to train a deep learning model sufficiently and achieve satisfactory performance.

To control the spread of the epidemic, lockdown measures have been proven to be effective^4,5,10^. After the lockdown starts, its influence on the spread of the COVID-19 will change over time. In particular, during the first several days of lockdown, the trend of the epedemic may not change because of the existence of the incubation, after which the change will take place and become more and more obvious^27^. This means that different attentions should be paid to the lockdown at different time points. However, few prediction models have considered such time-varying impact of the lockdown, which results in poor performance.

Our study in this paper is focusing on predicitng the epidemic spread trends in different countries with the governments switching the lockdown measurements. Specifically, we developed an attention-based RNN framework named ALeRT-COVID with transfer learning to to achieve the goal. We used the countries with rich data to build a source model (Fig. 2a) and transferred it to other target countries (Fig. 2b). Therefore, the prior knowledge learned from the source countries is transferred to complement the prediction for target countries. The attention mechanism was utilized to learn the time-varying impact of the previous case numbers on the final prediction. We evaluated the proposed model using mean absolute percentage error (MAPE) (Fig. 2c) and the results demonstrated our method can make more accurate prediction compared with other baseline models. Our work can help different countries to make decisions on whether the lockdown could be lifted or extended (Fig. 2d).

**Figure 2.**
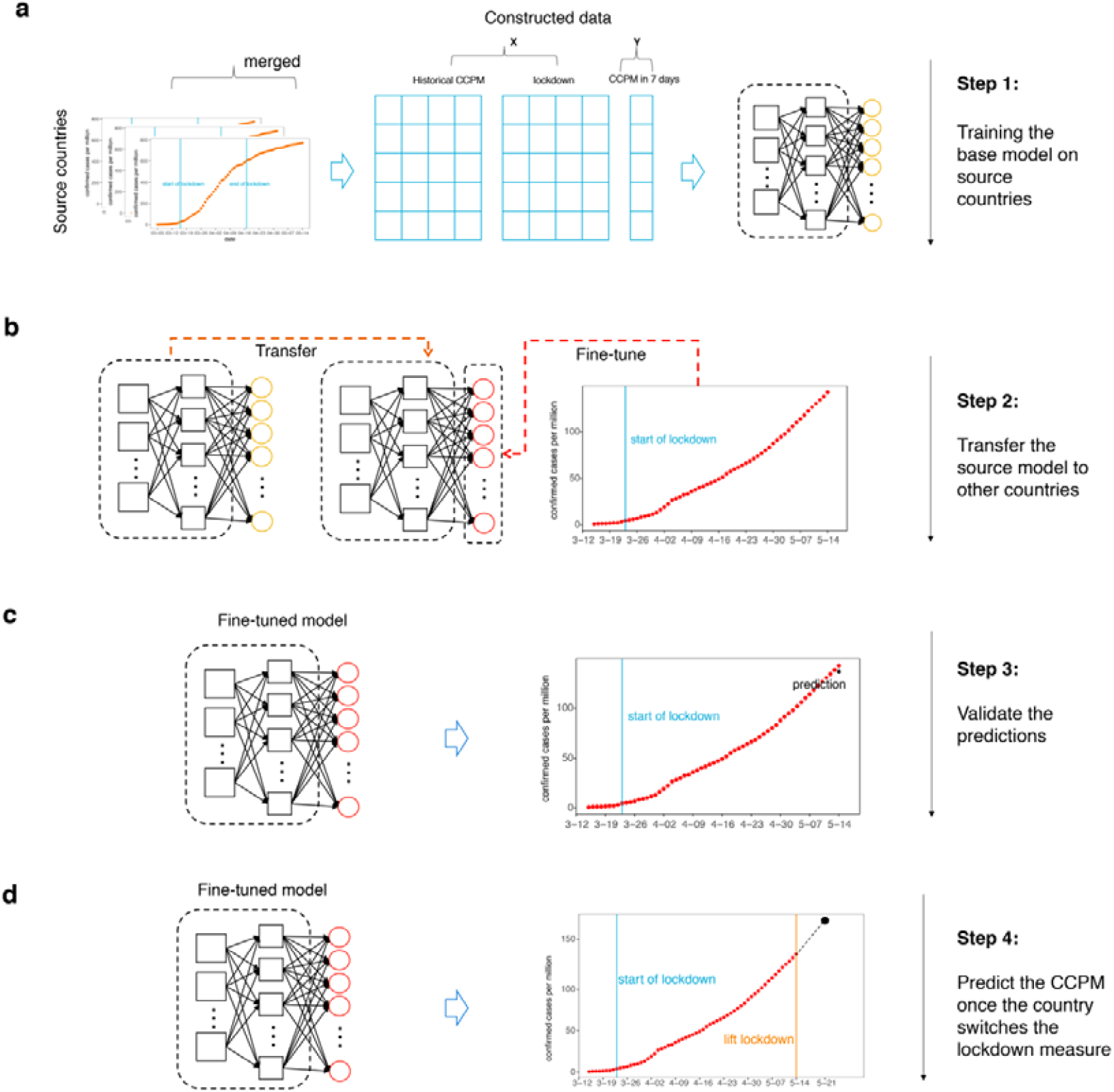
Schematic representation of the computational steps of ALeRT-COVID. **a**, The source model was trained on the source set constructed from the source countries. **b**, The source model was fine-tuned on each training set contructed from the correspomding target country. **c**, The fine-tuned model (target model) was validated on each test set contructed from the correspomding target country. **d**, The target model was utilized to predict the future CCPM (Confirmed Cases per Million) for the countries once the goverments switch the lockdown measures.

## Result

### Data Description

The data sources used in our study include the daily reported COVID-19 cases^28^ (update on May 31), the lockdown timeline^29^ and populations^30^ of 83 countries all over the world. These data were last updated on May 31, 2020. As the population sizes in different countries vary by a large range, we used the confirmed cases per million people (CCPM) in each country to estimate the severity of the epidemic. The date when the cumulative CCPM number reached one was used as the starting date for each country in our experiment. As the trends of Hubei and other parts of China displayed very different patterns, we treated them as two different areas in our data. For the countries reporting negative new daily cases due to the updated diagnositic critiera, we flatten their curves by using the average number of the neigbouring days.

The timeline of the COVID-19 progression for the 83 countries is illustrated in Fig. 1. As of May 31, the days since these countries announced the first COVID-19 cases range from 60 to 152 with an average number of 98. These countries implemented the lockdown measure at different periods, ranging from −7 to 82 with an average value of 27 days from the date of the first case in each country. Specially, the negative value −7 is corresponding to El Salvador, which started lockdown before the start of the epidemic. For the countries which had lifted lockdown as of May 31, the average length of the lockdown period is 47 days (min: 4 days; max: 127 days). Twenty countries are still under lockdown by May 31, among which 17 countries had not announced the date for lifting the lockdown.

The source countries are defined in our study as they have suffered from the epidemic for more than 84 days and lifted lockdown for more than 14 days. Finally, twelve trends of eleven countries, including Austria, China (except Hubei province), Hubei province, Croatia, Germany, Italy, Japan, Lebanon, Monaco, Norway, Oman and United Arab Emirates meet this criteria. Then the source set was constructed from these source countries and the 72 target sets are constructed from the other 72 countries (detailed in methods).

### Overall Predictive Performance of ALeRT-COVID

ALeRT-COVID first built the source model based on the CCPM sequences constructed from the source countries (detailed in Methods). Then the source model was transferred to each target country through a fine-tuning process. In order to evaluate the benefits of the three components in ALeRT-COVID, which are and 1) transfer learning 2) the lockdown information, and 3) attention mechanism, we also developed four baseline models for each target country.

- Linear regression model: A linear regression model with 7 predictors taking the cumulative CCPM values of the previous 7 days as input.
- Model A: using only the previous cumulative CCPM (confirmed cases per million people) as the predictor and training it directly on each country without transfer learning;
- Model B: using only the previous cumulative CCPM as predictor and fine tune the transferred source model;
- Model C: add the lockdown measure as an additional predictor for model B;

Mean absolute percentage error (MAPE) was utilized to evaluate the prediction performance. The most recent 20% cumulative CCPM numbers of each target country was used as the test sets to calculate the MAPE. Table 1 listed the overall MAPE scores on the 72 target countries. The mean MAPE was 0.118 (s.d. = 0.298) for the linear regression model. The standard deviation was large because the COVID-19 dispersions of some countries went far away from the linear pattern, which disagreed with the assumption of linear regression. The linear regression model therefore performed significantly worse for these countries. The mean MAPE was 0.100 (s.d. = 0.087) for model A (no transfer). The models using transfer learning (B, C and ALeRT-COVID) achieved improved predictive accuracies compared to model A. Among them, ALeRT-COVID, achieved the least mean MAPE score of 0.050 (s.d. = 0.040). Among all 72 target countries, 88.9% obtained MAPE scores less than 0.1 by ALeRT-COVID. These results demonstrated that the three components in ALeRT-COVID all helped enhance the prediction performance.

**Table 1.**
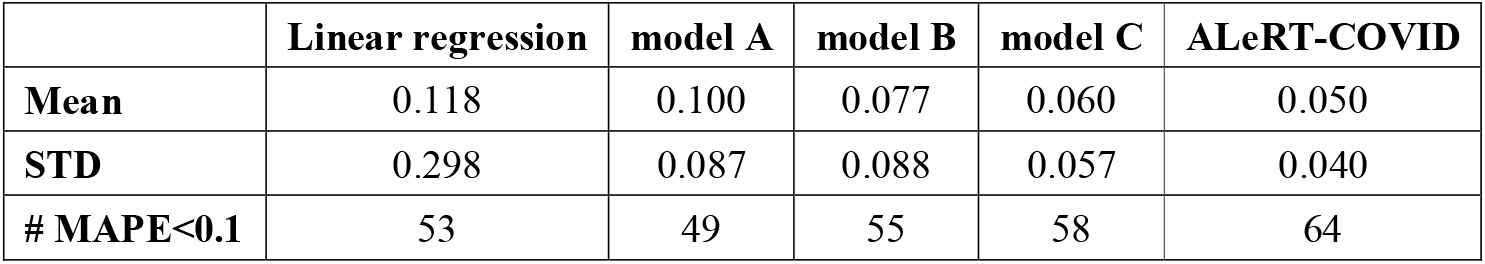
Summary Statsitics of MAPE on the test sets of 72 target countries by different models

|  | **Linear regression** | **model A** | **model B** | **model C** | **ALeRT-COVID** |
| --- | --- | --- | --- | --- | --- |
| **Mean** | 0.118 | 0.100 | 0.077 | 0.060 | 0.050 |
| **STD** | 0.298 | 0.087 | 0.088 | 0.057 | 0.040 |
| **# MAPE<0.1** | 53 | 49 | 55 | 58 | 64 |

### Prediction accuracies for countries lifting lockdown at different time

We selected three representative countries which lifted lockdown at different time periods to further demonstrate the effectiveness of adding the lockdown variable into the trend prediction model (Fig. 3). For Netherlands, their authorities have announced easing lockdown from May 19 (Fig. 3a), ALeRT-COVID yielded the best MAPE (0.007) compared to the other three models (model A: 0.020; model B: 0.02; model C: 0.033) on the test set. The predicted cumulative CCPM on May 31 by the model A (no transfer) and B (transfer) was much smaller than the ground truth. This was mostly because the model A and B did not consider lockdown and thus could not deal with the situation once the lockdown measure changes. In contrast, ALeRT-COVID and model C have incorporated the lockdown information, and thus produced much more precise predictions. Furthermore, ALeRT-COVID can predict better than model C, which suggests the benefits from the attention mechanism. For Singapore (Fig. 3b), although it announced lifting lockdown on the June 1, one day after May 31, ALeRT-COVID still yielded the best MAPE (0.034) compared to the other three models (model A: 0.259; model B: 0.062; model C: 0.058) on the test set. These result suggested that ALeRT-COVID could capture the pattern of the CCPM trend more precisely for countries lifting lockdown at different time periods.

**Fig 3.**
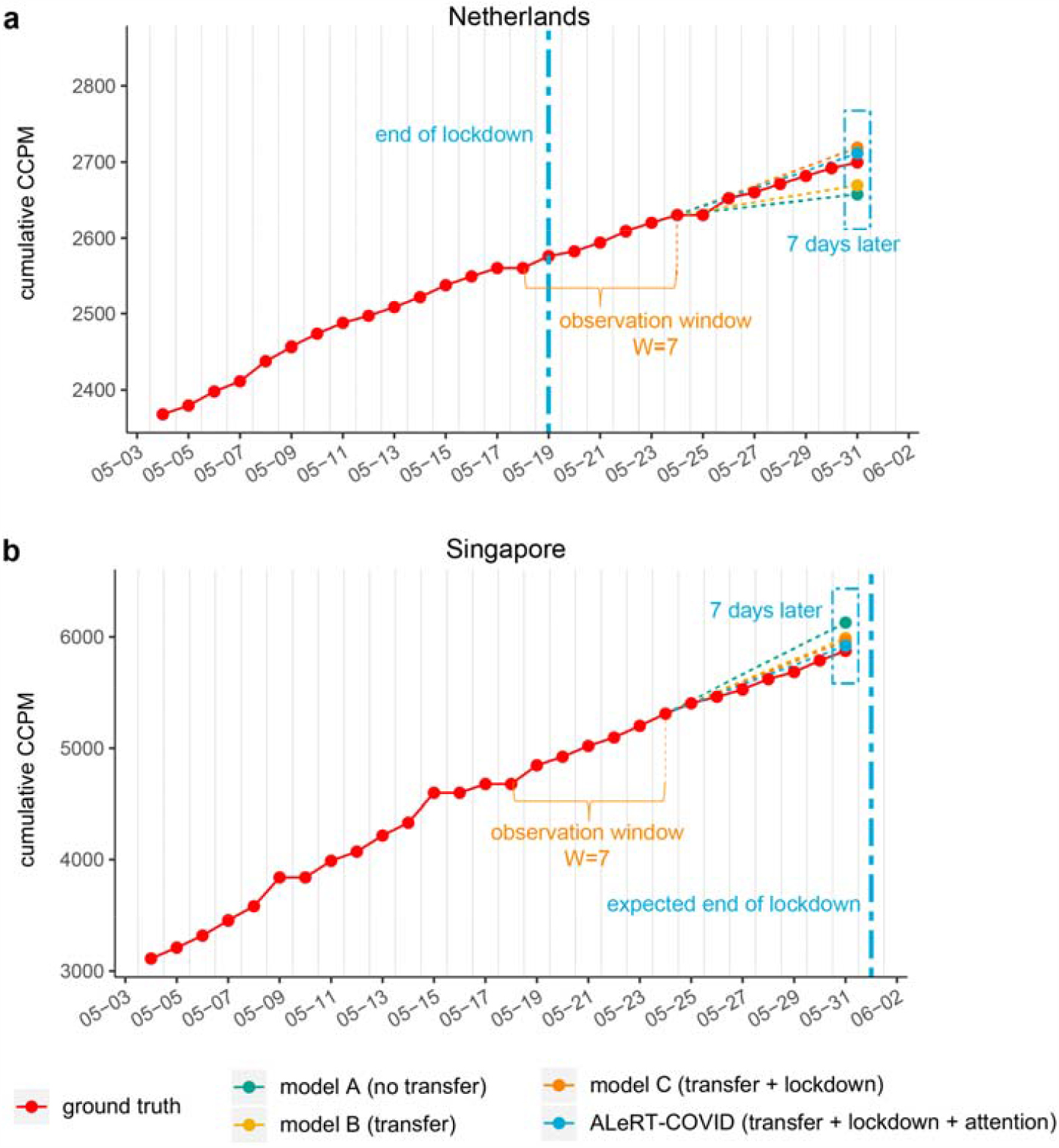
Prediction of the cumulative CCPM on May 31 for two representative countries that had announced the final date of the lockdown measure. The red points were the true cumulative CCPM on each day. The dashed vertical line represented the end date of the lockdown. The cumulative CCPM values and the lockdown information during May 18 to May 24 were input to the fine-tuned target model to predict the cumulative CCPM in seven days (May 31). The green, yellow, orange and blue points denoted the predictions of four different models.

### Predicting the change of the new case number when the lockdown measure is switched

We also used ALeRT-COVID to estimate the change of the new case number within 7 days by switching the lockdown measures. Specifically, for each specific country, we predicted two new case number during the period between June 1 to June 7: one was based on the actual lockdown measure (*pred* _*actual*_) and the other was based on the switched lockdown measure (*pred* _*switched*_). The change of the new case number within 7 days by switching the lockdown measures was calculated as

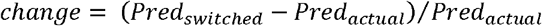

We chose two kinds of countries to do this analysis. The first group was composed of the countries that were still under lockdown until May 31 (Fig. 4a,b). The second group consisted of the countries that had lifted lockdown before May 25(Fig. 4c,d). The result demonstrated that the new case number from June 1 to June 7 of these target countries would increase or decrease to different extents.

**Fig 4.**
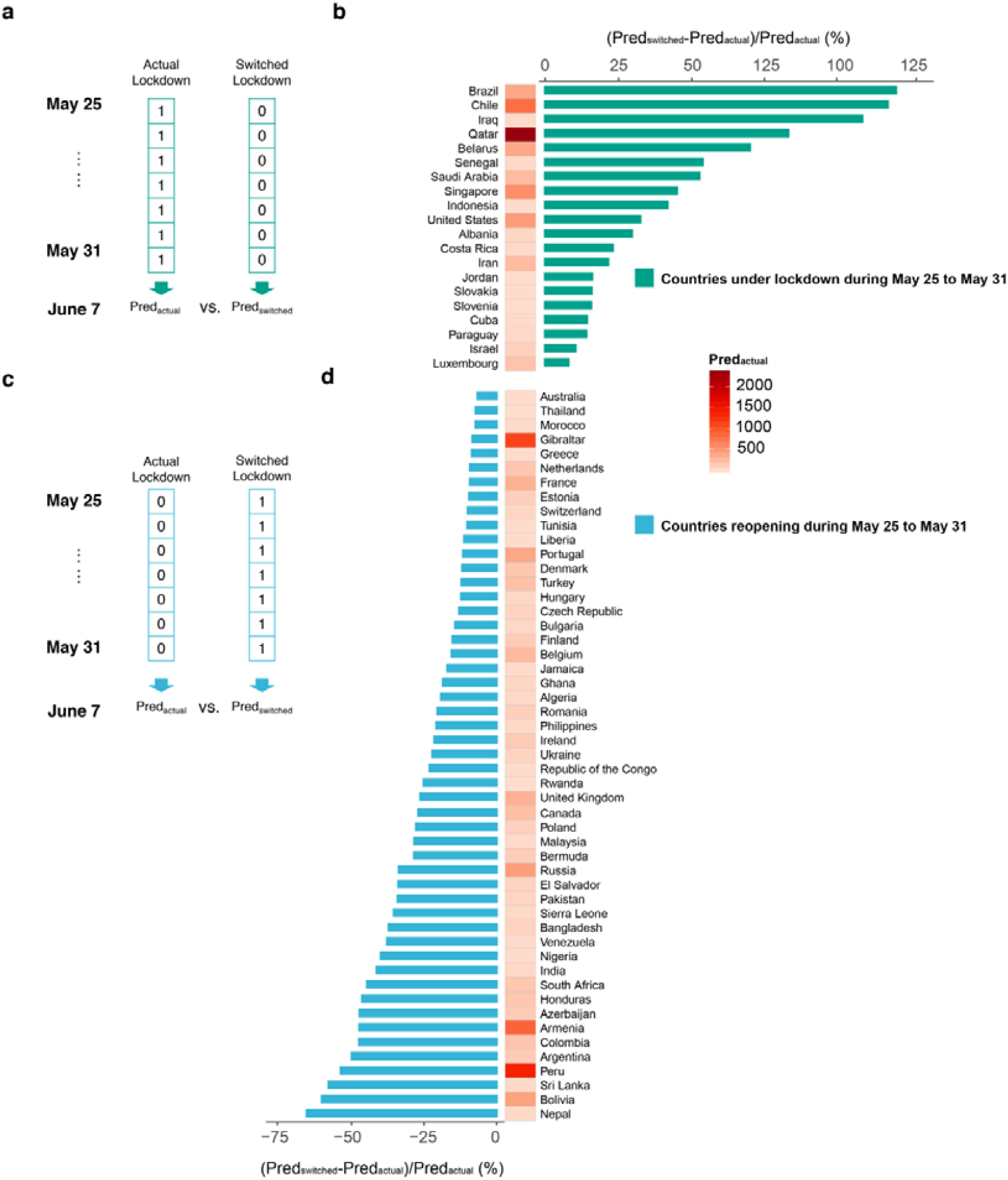
Change of the new case number in one week brought by switching the lockdown measure. **a)** Increasing percentage of the new case number in one week (June 1 to June 7) brought by lifting lockdown from May 25. **b)** Decreasing percentage of the new case number in one week (June 1 to June 7) brought by keeping lockdown during May 25 to May 31. The color of the cell in the heatmap represented the predicted new CCPM during June 1 to June 7 by ALeRT-COVID given the actual lockdown information. (*pred*_*actual*_ : predicted new case number based on the actual lockdown; *pred*_*switched*_ : predicted new case number based on switched lockdown;)

For those still under lockdown (Fig. 4b), once reopening, the new case number would raise. Specially, Brazil and Chile may suffer a doubled number of new cases in one week once the lockdown was lifted. This was mostly because these countries were in the rapid-growth phase of the epidemic (Fig. 5a,b). Lockdown measure took a crucial effect on controlling the epidemic progression. In constrast, lifting lockdown took moderate effect on some countries such as Israel and Luxembourg (Fig. 5c,d), because the new case number per day of these countries had decreased for a while.

**Fig 5.**
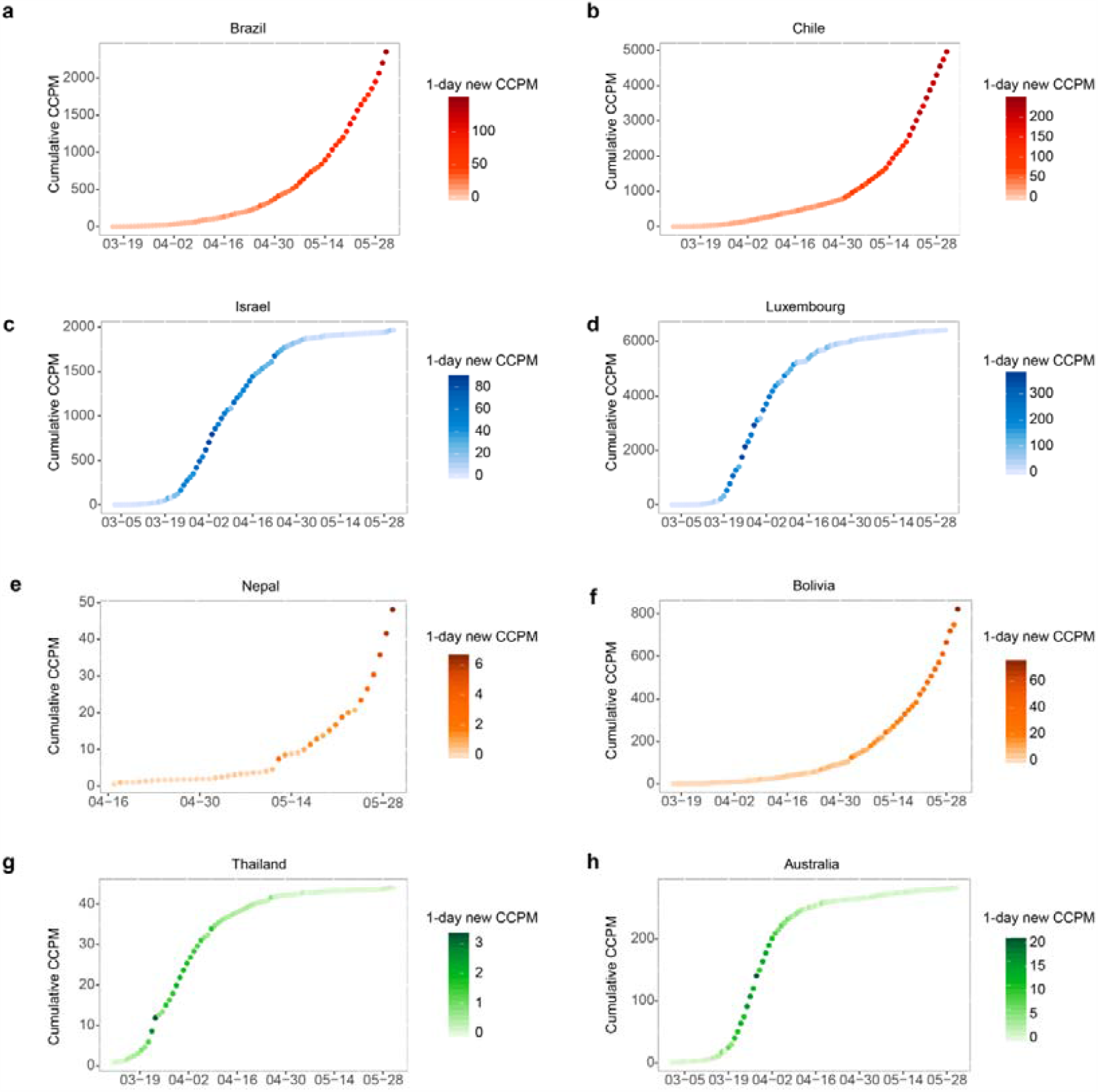
Cumulative CCPM Curve of eight countries. **(a,b,c,d)** Brazil, Chile, Israel and Luxembourg, which were under lockdown during May 25 to May 31. **(e**,**f**,**g**,**h)** Nepal, Bolivia, Thailand, Australia, which reopened during May 25 to May 31. Darker color of the point denoted larger new CCPM per day.

For countries that had already lifted lockdown before May 25 (Fig. 4d), resuming or extending the lockdown measure could reduce the new case number. Particularly, the new case number of Nepal and Boliva between June 1 to June 7 might decrease by half if they remained lockdown. This is mostly because the cumulative CCPM of both countries continued steady increase (Fig. 5e,f). Lockdown measure still helped to large extent in controlling the spread of the COVID-19. On the contrary, for countries like Australia and Thailand (Fig. 5g,h), as the new case per day was decreasing, there were no big difference between extending and lifting the lockdown in our predictions.

## Discussion

We proposed a deep learning approach named ALeRT-COVID to predict the COVID-19 spread trend of different countries around the globe. There were three main novelties in our approach. First, we utilized transfer learning^31^ to deal with the insufficient-data problem of the countries in the early-stage of the COVID-19. Second, we added the lockdown information into the predictors of the proposed model. This enabled ALeRT-COVID to capture the effect of the lockdown measure and more precisely forcast the change of the future case number when the lockdown measure switches. Third, ALeRT-COVID utilized the attention mechanism^32^ to capture the contribution of previous CCPM numbers on the trend, which greatly enhanced the model as reflected on the improvement of its prediction performance. In the following we expand these three points with more details.

Transfer learning^31^ can help enhance the deep learning based prediction models especially for countries without sufficient data due to the early-stage epidemic. There have been existing studies applying deep learning algorithms to predict the epidemic progression in specific countries^15,21,33^. However, they all trained the model directly on the data from these countries. Although deep learning models can achieve much more accurate results, they often require a large amount of data points to train the model, which is difficult for the early-stage countries who just started the COVID-19 crisis for a short time. In contrast, transfer learning mechanisms built the source model based on the source countries with enough training points and then fine-tuned its parameters to better fit the specific target country, which greatly reduced the amount of required training data samples from the target countries with the prior knowledge learned from the source countries^31,34^. As an example in our results, among the early-stage countries, Sierra Leone underwent the epidemic for 52 days from the date when the cumulative CCPM reached 1, with inadequate samples in the training set. It gained a MAPE score of only 0.202 for models without transfer learning, while transfer learning improved the performance significantly (model B: 0.058; model C: 0.047; ALeRT-COVID: 0.041).

Lockdown measures have been proved to play an important role in controlling the rapid growth of CCPM ^8,35,36^ Adding the lockdown information could boost the model performance especially for the time points after lifting the lockdown measures. As illustrated in our results, model A and B, which did not use the lockdown information, predicted significantly lower CCPM values than the ground truth for Canada and Netherlands after they lifted the lockdown measure on May 4 and May 19 respectively (Fig. 3a, b). As most of the training data of model A and B was from the previous CCPMs under the lockdown, the model trained on them were unable to learn the logical trend when lockdown has been lifted. Consequently, their predictions still followed a slow growth course even the country has lifted lockdown already for more than ten days. In contrast, Model C and ALeRT-COVID, which incorporated the lockdown infromation could produce much closer predictions to the ground truth.

In addition, ALeRT-COVID added the attention mechanism to better capture the contribution of the previous CCPM numbers on the future trend. In our expereriments, we utilized the cumulative CCPM of the last seven days to predict the CCPM in one week. In real world situations, the previous case number and the lockdown measure in the past days have different impact on the epidemic progression. Without the attention mechanism, the previous case of each day are assumed to have the same effect on the future trend, which may not be true. The attention mechanism allows these previous case numbers to have differential impact on the future predictions, which makes the model more flexible and thus leads to more accurate predictions.

More importantly, having the lockdown measure can also help predict the impact on the epidemic progression by changing such control policy. We used ALeRT-COVID to predict the new CCPM for certain countries from June 1 to June 7 supposing if they have released the lockdown (still under lockdown in fact) or extended lockdown (reopening already in fact) during May 25 to May 31 respectively.

Once lockdowns have been lifted, we found that the new case number in one week would increase significantly for countries such as Brazil and Chile. They would suffer a double number of new cases in 7 days once the lockdown was lifted. This is mostly because the epidemics in these countries are still in rapid growth. Because of the critical role of lockdown in controling the spread of COVID-19, these countries should consider extending their lockdown measure for a longer time. In contrast, lifting lockdown would not cause a significant increase to the COVID-19 spread for countries like Israel and Luxembourg because the epidemics in these countries had already entered into a gentle phase. In other words, the number of new cases per day had been decreasing for a while. It is safer for these countries to consider releasing the lockdown measure shortly to recover the economy. Similar patterns were presented in the countries which had lifted the lockdown as of May 25. For countries like Nepal and Bolivia, although they were still undergoing a fast increase of the number of cases, the lockdown had been lifted on May 7 and May 10 respectively. From our prediction, the number of new cases within seven days could be reduced to about fifty percent if these countries remained lockdown. For countries such as Thailand and Australia, whose COVID-19 spread trends had already entered into a gentle phase, showed no significant decrease of the new case number when extending the lockdown measure. Therefore, our results indicate that it is important to keep the lockdown for the countries that still undergoes a rapid gowth of the case number, it may be a better choice to extend or resume the lockdown measure, and it is relatively safer to release the lockdown measure for the sake of economy recovery for the countries with decreasing number of new cases.

There are several limitations of our research. First, the lockdown measure was modeled as a binary variable representing the lockdown is on or off. However, different countries conducted different levels of the lockdown. Even within the same country, such as United States, different states had different lockdown measures. A more fine-grained quantification of the lockdown measures may enhance the prediction performance. In addition, more related information could be introduced to the model such as the number of recovered cases, death cases, available healthcare resources, etc. Although the proposed method aims at predicting the cumulative case numbers, the prediction targets can be conveniently changed to the number of recovery or death cases.

In conclusion, our predictions provide valuable data-driven insights for better understanding the current situation and could help policy makers and health authorities make plans to manage the future situation.

## Methods

### Constructing source and target sets

We denote the orginal data set associated with each specific country *i* as, *C* stands for the cumulative CCPM data and *L* represents the lockdown data.,. corresponds to the date when the cumulative CCPM number reached 1. equals to the number of days from date of t=1 to May 31 in our experiment. Given an observation window W and a prediction window P, *ALeRT-COVID* takes as input and predict the output value,. We construct} for each specific country. Specifically, we set both the W and P to 7 days in our paper but they can be changed to other values.

The source set is obtained by merging the from all source countries or area (Austria, China (except Hubei province), Hubei province, Croatia, Germany, Italy, Japan, Lebanon, Monaco, Norway, Oman and United Arab Emirates). The target set is constructed from the target country. Finally, we construct one source set composed of 893 samples and 72 target sets consisting of different number of samples ranging from 32 to 104.

### Architecture of ALeRT-COVID

The architecture of *ALeRT-COVID* is illustrated in Figure 6. There are two main components in *ALeRT-COVID*, aiming to encode the previous CCPM sequence and incorporate the effect of the lockdown measure into the prediction respectively. In the following descriptions, we omit the country index *i* as the superscript for notation simplification.

**Figure 6.**
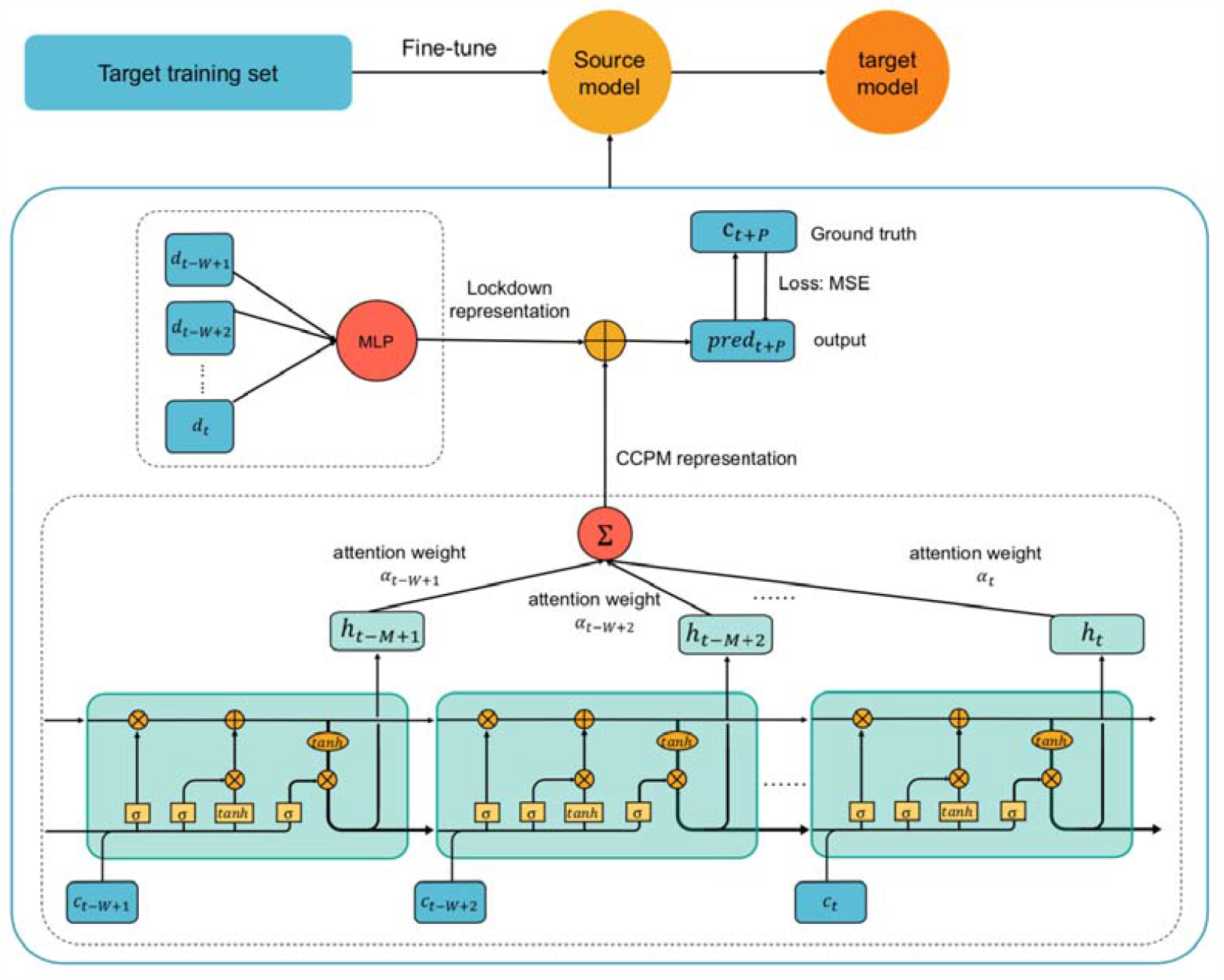
Architecture of ALeRT-COVID.

The standard Long-Short-Term-Memory (LSTM)^37^ is employed to encode the CCPM sequence data. More specifically, LSTM generates hidden states *h*_*τ*_ based on the history *C*_(t –W, *τ*]_ for any *τ* ∈(t − *W*,t], where *t* ∈(*W,T*_*max*_ − *P*]. The hidden states *H*_*(t − w,t*]_, are then sent into MLP to learn the attention^32^ weights *α* _*(t* − *W*,t]_, for identifying which time steps influence most on the final prediction. The encoded CCPM sequence is thus obtained by ∑_*τ ∈(t − W,t*]_ (α _*τ*_ ×*h*_τ_).

The effect of the lockdown measure on the final prediction is learnt by an MLP taking both the lockdown sequence *L* _*(t* − *W,t*]_,, and the CCPM sequence *C* _*(t* − *W,t*]_,, together as input. The output of the MLP is denoted as *g*_*t*_.

Then the final output is ∑_*τ* ∈(t − *W*,t]_ (α _*τ*_ ×*h*_τ_)+*g*_*t*_ We use the mean square error (MSE) to calculate the final loss.

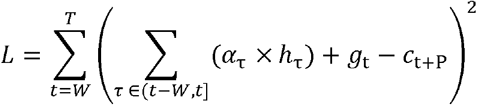

where *T*=*T*_*max*_− *P*.

### Model configurations and Training details

#### Source model

Given the observation window W=7 days, we train an LSTM of length 7. We explored different hyperparameters. We tried different number of LSTM layers from {1, 2, 3}. We also explored different numbers of nodes in the LSTM cell that gradually increases from 2,4 to 10. The activation functions of the LSTM include hyperbolic tangent (tanh) and rectified linear unit (ReLU). The number of attention layers is set to 1. The activation function is softmax. The number of layers of the MLP for lockdown effect is set to 1 and the activation function is selected from {tanh, ReLU}. Particularly, we use the “weight constraints” in Keras^38^ to force the weights corresponding to the lockdown vector to be non-positive. We do this because the positive weight of the lockdown vector demonstrates that implementing the lockdown measure will raise the future CCPM number, which is unlikely to happen in reality. The optimizers include ADAM^39^, Stochastic Gradient Descent^40^and RMSProp^41^. The batch size varies from 16, 32, and 64. For the source model, we use 5-fold cross validation on the source set to find the optimal hyperparameters from the above settings. Finally, the source model uses ADAM optimizer, ReLU activation in LSTM, ReLU activation in MLP for lockdown effect and batch size of 32.

#### Target model

Each of the target set is split into a training part (first 80%) and a validation part (last 20%). After training the source model on the source set, we adapt it to each target training set. The architecture of the source model remains the same during transfer learning. We freeze the LSTM parameters in the source model and re-trained (fine-tuned) other paramters in the MLP for lockdown effect and the attention layers on the training part of. We use the ADAM optimizer and batch size of 4.

#### Baseline Models

Here we present the architecture of the baseline models we compared in our experiments.

- Model A: LSTM with a fully-connected layer as the output layer. The architecture of the LSTM is the same with that in *ALeRT-COVID*. The input of model A for a specific country *i* contains only the CCPM sequence *C*^*i*^ _*(t* − W,t]_. The model is trained directly on the training part of the target set without transfer learning.
- Model B: LSTM with a fully-connected layer as the output layer. The architecture of the LSTM is the same with that in *ALeRT-COVID*. The input of model B for a specific country *i* contains only the CCPM sequence *C*^*i*^ _*(t* − W,*t*]_. Transfer learning is utilized. A source model is constructed first on the source set and an target model is obtained by fine-tuning the parameters of the last fully-connected layer on the training part of the target set.
- Model C: LSTM with a fully-connected layer as the output layer. The architecture of the LSTM is the same with that in *ALeRT-COVID*. The input of model C for a specific country *i* contains both the CCPM sequence and the {*C*^*i*^ _*(t* − W,*t*],_ *L*^*i*^_*(t* − W,*t*]_}. Transfer learning is utilized. A source model is constructed first on the source set and an target model is obtained by fine-tuning the parameters of the last fully-connected layer on the training part of the target set.

#### Model Evaluation

All the compared models are evaluated on the validation part of each target set by the mean absolute percentage error (MAPE), which is defined as follows,

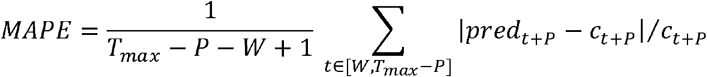

where *pred*_*t +p*_ is the predicted cumulative CCPM at the time point t+P,*c*_*t +p*_ is the ground truth.

## Data Availability

The data can be obtained from the offered links or upon request to the authors.

https://auravision.ai/covid19-lockdown-tracker/

https://en.wikipedia.org/w/index.php?title=List_of_countries_and_dependencies_by_population&oldid=960653268

https://www.worldometers.info/coronavirus/

## Data & Code availability

Data & Code for the ALeRT-COVID model is available at https://github.com/wcmwanglab/ALeRT-COVID.The data sets of all the source countries and several sample target countries can be obtained from the GitHub link. All other data may be obtained upon request to the authors or downloaded from https://www.worldometers.info/coronavirus/#countries.

## Notes

### Competing Interest Statement

The authors have declared no competing interest.

### Clinical Trial

n/a

### Funding Statement

no external funding was received

### Author Declarations

Our study does not use any patient level data and is purely epidemiological study with publicly available regional data

